# Immunization Opportunities for Hospitalized Adolescents

**DOI:** 10.1101/2022.09.10.22279805

**Authors:** William R. Wurster, Byron A. Foster, James M. Walston, Tiffany A. Gardner, Hanae Benchbani, Jared Austin

**Affiliations:** Doernbecher Children’s Hospital, Portland, OR; Oregon Health and Science University, Portland, OR

**Author notes:** **Address correspondence to:** William R. Wurster, MD, Doernbecher Children’s Hospital, Oregon Health and Science University, 700 SW Campus Drive, Portland, OR 97239, [ ].

## Abstract

**Introduction:** Adolescents seek routine healthcare, including immunizations, less frequently than any other age group. Hospitalizations are an opportunity to provide immunizations to this vulnerable population. The aims of this study were to assess the accuracy of provider documentation of immunization status and evaluate the prevalence of delayed immunization status in this population.

**Methods:** A retrospective chart review of adolescents discharged from July 2017 to June 2018 from the pediatric hospitalist service of a tertiary care academic children’s hospital was conducted. Provider documentation of immunization status was compared to the immunization registry ALERT Immunization Information System (ALERT IIS) linked to the electronic medical record using descriptive statistics.

**Results:** Provider documentation of up-to-date on all immunizations had a sensitivity of 60% and specificity of 55%, with 84% of patients needing at least one immunization despite 48% of patients being documented as up-to-date by providers. Provider documentation of the immunization status for the HPV, MCV and Tdap immunizations displayed a low sensitivity (10-11%) but a high specificity (97-100%) while documentation of the influenza immunization was associated with high sensitivity (86%) and low specificity (26%). Provider documentation of immunization status for the HPV, MCV, Tdap and influenza immunizations had positive likelihood ratios of 3.5, 8.5, infinity, and 1.2 with negative likelihood ratios of 0.9, 0.9, 0.9 and 0.53, respectively.

**Conclusions:** Providers inaccurately documented the immunization status for adolescent patients in the inpatient setting. Hospitalizations may provide opportunities to improve immunization rates in adolescents, especially when using state immunization registries.

## Introduction

Immunizations are a critical component of adolescent preventive medicine, and the adolescent population has increasingly been recognized as a source for multiple immunization preventable diseases.^1^ Unfortunately, adolescents have some of the lowest immunization rates of any age group, and only 48% of adolescents aged 13-15 years received the recommended HPV immunization doses by 2018 which is far from the Healthy People 2030 objective of 80%.^2^ Therefore, a more comprehensive strategy for delivering immunizations to adolescents is needed, including the delivery of immunizations in non-traditional times and settings, such as during hospitalization.

Over 5 million children and over 700,000 adolescents are hospitalized each year and this presents a unique opportunity to provide preventive care services, including immunizations, to this vulnerable population.^3,4^ Indeed, two recent studies in the United States evaluated the immunization status of inpatient pediatric patients of all ages and found that adolescents accounted for the majority of those under immunized.^5,6^ To address this, multiple interventions to improve immunization rates for hospitalized pediatric patients have been studied, varying from simply identifying the under immunized and offering immunizations to implementing complete immunization teams.^5,7-9^ Electronic medical record (EMR) reminders, increasing nursing participation, creating clinical practice guidelines and adding “under immunization” to the inpatient problem list have been shown to improve the immunization rates of hospitalized pediatric patients.^7,10,11^

One challenge to providing immunizations to hospitalized adolescents is obtaining accurate immunization records.^7^ Multiple studies have assessed validity of parental recall compared to medical records and found that parental recall overestimates or underestimates the immunization status of pediatric patients.^5,7,12-16^ Accuracy of provider documented immunization status in the EMR is another challenge to providing immunizations to adolescents.^17^ One study found that Pediatric Emergency Department and inpatient providers documented 92% of pediatric patients presenting for an acute respiratory illness up-to-date on their immunizations while only to 42% of those patients were documented as up-to-date in the immunization registry.^18^ Another study found that accuracy of immunization status documentation for inpatient adolescents did not change despite modifying discharge order sets in the EMR and providing training seminars to providers about the administration of the HPV immunization.^19^

Our objectives were to investigate the accuracy of provider documentation of immunization status of hospitalized adolescents and evaluate the prevalence of delayed immunization status among this population.

## Methods

### Study Setting/Participants

A retrospective chart review was performed from July 2017 to June 2018 of adolescent patients (aged 11-19 years) discharged from the pediatric hospitalist service at a 150-bed tertiary care academic medical center located in the Pacific Northwest. All documentation was present in the electronic medical record (EMR) and care for all patients was provided by resident and attending physicians. No inpatient program existed to provide adolescents immunizations.

### Inclusion and Exclusion criteria

All adolescent patients discharged from the pediatric hospitalist service from July 2017 to June 2018 were included in the study. This population consisted of patients admitted to the hospitalist service from an outside hospital ward, ICU or emergency department, and those patients transferred from the PICU or admitted from the emergency department within our own institution. In order to screen out patients who may have received immunizations in other states, patients needed at least one non-influenza immunization documented in the immunization registry to be included. If the patient had multiple admissions over the study time frame, the data from the first admission was only included in the analysis. No patients were excluded based on diagnosis. Participants were excluded if their primary home address in the EMR was outside the state of Oregon and if they did not have at least one non-influenza immunizations documented in the ALERT Immunization Information System in the EMR. In the state of Oregon, the ALERT Immunization Information System (ALERT IIS) is used to track patient immunization status through the use of medical records. ALERT IIS is a lifetime, statewide immunization registry that encompasses patients of all ages and consolidates immunization reports into a single record for each patient. All pediatric clinics, family medicine clinics, pharmacies, rural health clinics, migrant health centers, local health department clinics, federally qualified health centers and school-based health centers in the state of Oregon report to ALERT IIS. All providers participating in the “Vaccines for Children” program in the State of Oregon are required by the CDC to report all immunizations to ALERT IIS within 14 days.^20^

### Data Collection

Patients discharged from July 2017 to June 2018 were identified from the billing database of the hospital. The principal investigator trained two research assistants to be the primary data extractors. The research assistants would flag charts for review by the principal investigator if they had questions regarding data collection. Data were collected and entered into a secure REDCap database.

Data were collected on immunizations recommended for the adolescent population: quadrivalent meningococcal (MenACWY or MCV), tetanus and reduced diphtheria toxoids and acellular pertussis (Tdap), human papillomavirus (HPV) and influenza.^21^ The admission history and physical, transfer note (if applicable) and discharge summary were reviewed for provider documentation of immunization status. The admission history and physical note template included a section to free text immunization status while the transfer and discharge summary note templates did not include a dedicated section for immunization status. Resident physicians completed the majority of history and physical, transfer and discharge summary notes. The ACIP guidelines for adolescent vaccines and the patient’s age on the date of admission were used to determine the patient’s status for each vaccine using the ALERT IIS.^22^ The ALERT IIS immunization record is embedded in the EMR under an “Immunizations” tab in each patient’s chart. A patient was only considered overdue for the influenza immunization when it was available for administration from September 2017 to April 2018. The total number of non-influenza immunizations documented in ALERT IIS was also obtained from the EMR to serve as a marker for overall immunization history.

### Dependent Variables

The primary outcome of interest was the concordance between provider documentation of immunization status and the immunization status present in the ALERT IIS. A secondary outcome of interest was the prevalence of delayed immunization status among the hospitalized adolescent population. Other outcomes of interest included the number of immunizations provided to this patient population during their hospitalizations.

### Independent Variables

Demographic data including age, gender, race, ethnicity, insurance type and technology dependence, and admission data including primary discharge diagnosis category, length of stay, admit or transfer information and gender of admitting resident were obtained from the EMR.

### Statistical Analysis

Immunization data were initially analyzed using descriptive statistics. Sensitivity, specificity, positive likelihood and negative likelihood ratios were calculated for each immunization using IBM SPSS Statistics.^23^ For these calculations, provider documentation of the patient needing the immunization was considered “positive test” while documentation of the patient being eligible to receive the immunization in the ALERT IIS was considered “disease positive.” No provider documentation of the patient needing the immunization was considered “negative test”. If the patient was up-to-date on the immunization in ALERT IIS, this was considered “disease negative.” Thirty-three percent of patients had provider documentation that the immunization status was unknown, or no immunization information was documented by a provider.

### Ethics

This study was deemed exempt by our Institutional Review Board. All charts were reviewed by study team members, and no personal health information was shared outside of the organization.

## Results

### Sample Characteristics

A total of 207 adolescent patients were discharged from the pediatric hospitalist service during the study period. After EMR review, 160 patients were included in primary statistical analysis (Figure 1). Overall, the average age was 14.4 years and 53% of patients identified as female (Table 1). The majority were white, non-Hispanic, with public health insurance. The most common primary discharge diagnosis category was intentional ingestion, and the average length of stay was roughly 5 days. After reviewing the ALERT IIS, patients up-to-date on their immunizations had longer hospital stays compared to patients needing catch up immunizations (mean 8.6 days vs 4.3 days, P = 0.003).

**Figure 1.**
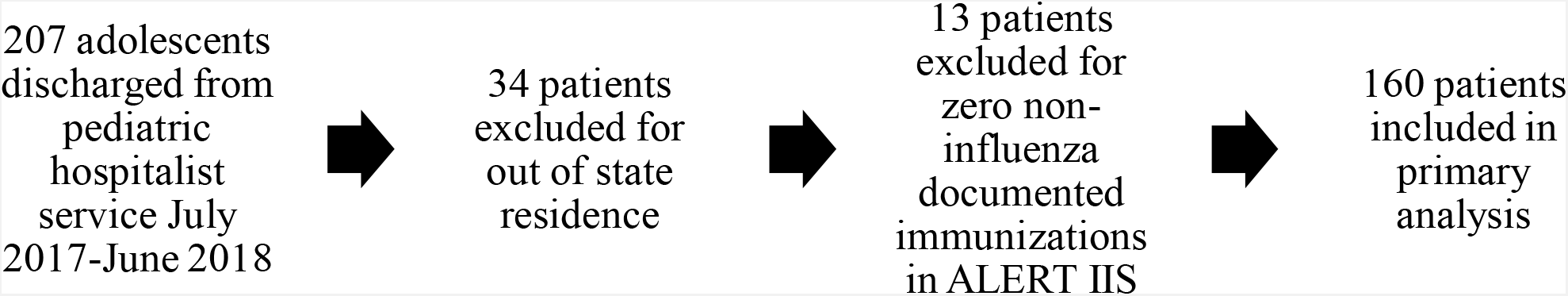
Flow diagram of inclusion and exclusion criteria.

**Table 1.** Demographic differences between patients up-to-date on their immunizations and those needing catch up immunizations based on review of the ALERT IIS in the EMR.

|  | Total Population:<br>160 patients | Up-to-date: 25 patients | Delayed: 135 patients |
| --- | --- | --- | --- |
| Age in years, mean (SD) | 14.4 (2.2) | 15.1 (2.1) | 14.4 (2.2) P=0.106 |
| Gender |  |  | P=0.477 |
| • Male | 72 (45) | 13/72 (18) | 59/72 (82) |
| • Female | 85 (53) | 12/85 (14) | 73/85 (86) |
| • Gender non-conforming | 3 (2) | 3 (100) | 0 |
| Race |  |  | P=0.822 |
| • White | 135 (84) | 21/135 (16) | 114/135 (84) |
| • Asian | 9 (6) | 2/9 | 7/9 |
| • Other | 16 (10) | 2/16 | 14/16 |
| Ethnicity |  |  | P=0.797 |
| • Hispanic | 28 (17) | 5/28 (18) | 23/28 (82) |
| • Non-Hispanic | 131 (82) | 20/131 (15) | 111/131 (85) |
| • unknown | 1 (1) | 0 | 1 |
| Insurance Type |  |  | P=0.379 |
| • Private | 71 (44) | 9/71 (13) | 62/71 (87) |
| • Public | 87 (55) | 15/87 (17) | 72/87 (83) |
| • Uninsured | 0 (0) | 0 | 0 |
| • unknown | 2 (1) | 1/2 | 1/2 |
| Medical technology dependence |  |  | P=0.183 |
| • Yes | 28 (18) | 8/34 | 26/34 |
| • No | 132 (82) | 17/126 | 109/126 |
| Primary discharge diagnosis category |  |  | P=0.111 |
| • Intentional Ingestion | 42 (26) | 6/42 (14) | 35/42 (86) |
| • Skin/MSK | 25 (16) | 5/25 (20) | 20/25 (80) |
| • Respiratory | 23 (14) | 3/23 (13) | 20/23 (87) |
| • Neurologic | 18 (11) | 6/18 (33) | 12/18 (66) |
| • GI | 16 (10) | 3/16 (19) | 13/16 (81) |
| • Psych | 12 (8) | 0/12 (0) | 12/12 (100) |
| • GU | 9 (6) | 0/9 (0) | 9/9 (100) |
| • other | 15 (9) | 3/15 (20) | 12/15 (80) |
| Admit vs Transfer to Peds Hospitalist group |  |  | P=0.803 |
| • Admit | 114 (71) | 18/114 | 96/114 |
| • Transfer | 46 (29) | 9/46 | 37/46 |
| Length of Stay in days, mean (SD) | 5.3 (7.7) | 8.6 (12.0) | 4.3 (5.1) P = 0.003 |
| Gender of admit resident |  |  | P=0.489 |
| • Male | 49 (31) | 6/49 | 43/49 |
| • Female | 111 (69) | 19/111 | 92/111 |

### Accuracy of Provider Documented Immunization Status

Provider documentation of immunization status underreported the need for immunizations when compared to the immunization registry ALERT IIS in the EMR. Providers documented that patients needed immunizations at the following rates: HPV 8%, MCV 6%, Tdap 2% and influenza 21% (Figure 2). On review of the ALERT IIS, 59%, 51%, 19% and 76% of patients needed the HPV, MCV, Tdap and influenza immunizations, respectively. Forty-eight percent of patients were documented by providers as up-to-date on their immunizations whereas 16% of patients were up-to-date in the ALERT IIS (Figure 2). The 95% confidence intervals for provider documented immunization status were statistically different compared with the 95% confidence intervals obtained from review of the ALERT IIS for all of the immunizations studied including when providers documented patients as up-to-date on their immunizations (Figure 2, Table 2).

**Figure 2.**
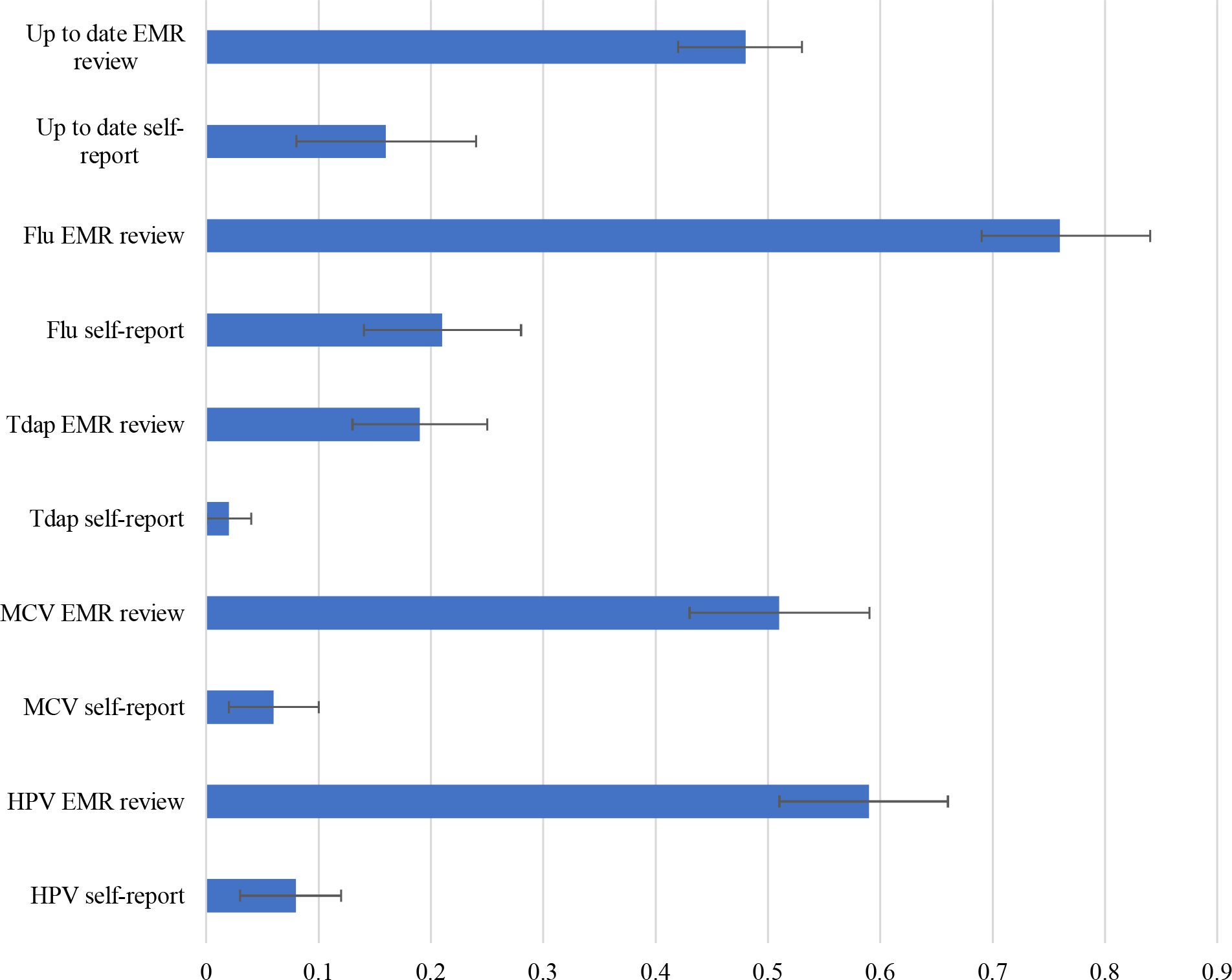
Parent/Patient self-report of immunization status compared to review of ALERT IIS

**Table 2.** Sensitivity, Specificity and Likelihood Ratios (LR) of Provider documentation of Immunization Status

| Immunization | Sensitivity | Specificity | Positive LR | Negative LR |
| --- | --- | --- | --- | --- |
| HPV | 11% | 97% | 3.5 | 0.9 |
| MCV | 11% | 99% | 8.5 | 0.9 |
| Tdap | 10% | 100% | ∞ | 0.9 |
| Influenza | 86% | 26% | 1.2 | 0.5 |
| Up to date | 60% | 55% | 1.3 | 0.7 |

HPV = human papilloma virus immunization
MCV = meningococcal immunization
Tdap = tetanus, diphtheria and acellular pertussis immunization
LR: likelihood ratio

Thirty-three percent of the patients did not have provider documentation of their immunization status, or the immunization status was documented as unknown in the history and physical, transfer or discharge summary notes.

Provider documentation of immunization status for the HPV, MCV and Tdap immunizations displayed a low sensitivity (10-11%) but a high specificity (97-100%) using the ALERT IIS system as the standard (Table 2). If the patient was due to receive the immunization in ALERT IIS, providers only documented 10-11% of those patients as due for the immunization in the history and physical, transfer or discharge summary notes. Conversely, provider documented immunization status for the influenza immunization was associated with a high sensitivity (86%) and a low specificity (26%) (Table 2). If the patient was eligible to receive the influenza immunization in the ALERT IIS, providers documented that 86% of those patients needed the immunization. Provider documentation of up-to-date immunization status had a sensitivity of 60% and specificity of 55%.

Provider documented immunization status for the HPV, MCV, Tdap and influenza immunizations had positive likelihood ratios (LR+) of 3.5, 8.5, infinity, and 1.2 with negative likelihood ratios (LR-) of 0.9, 0.9, 0.9 and 0.5, respectively (Table 2).

### Immunizations Given During Hospitalization

Of the 84% (135/160) of patients eligible to receive one or more immunizations in the ALERT IIS, 24% were provided immunizations prior to discharge. These 32 patients received 35 total immunizations: 3 HPV, 1 MCV, 1 Tdap, 28 influenza and 2 other immunizations. Of the patients eligible to receive the adolescent immunizations (HPV, MCV, Tdap) in the ALERT IIS, 1-3% of those patients received those immunizations prior to discharge. One patient received the HPV, MCV, Tdap and influenza immunizations. Of the 90 patients eligible to receive the influenza immunization in the EMR, 28 received influenza immunizations prior to discharge.

## Discussion

Our study illustrates that delayed immunizations are highly prevalent in hospitalized adolescents with 84% (135/160) of patients needing at least one immunization. Additionally, provider documentation of immunization status for hospitalized adolescent underreports the need for immunizations compared to the immunization registry ALERT IIS. This was true for all major recommended immunizations for this age group and for influenza. While our study illustrates the opportunity to improve the immunization rates of this population, few immunizations outside of seasonal influenza were actually provided.

Published literature illustrates that adolescents are particularly at risk for under immunization.^1,2,5,6^ Multiple studies evaluating pediatric patients of all ages found adolescents to account for the majority of patients under immunized and increasing age was a risk factor for being under immunized.^5,6^ Adolescents are less likely to have a medical home and seek routine health care less than any other age group, illustrating the importance of using every opportunity, including hospitalizations, to provide immunizations to this vulnerable population.^1,3,24^

Obtaining accurate immunization records is a barrier to providing immunizations in the inpatient setting.^5,7^ In this study, when comparing the immunization registry ALERT IIS to provider documented immunization status, overestimation of immunization status by providers was statistically significant for all immunizations investigated. Provider documented immunization status for the three immunizations primarily administered during adolescence (HPV, MCV, Tdap) had a low sensitivity and high specificity. In contrast, provider documented immunization status for the influenza immunization had a high sensitivity and a low specificity. In addition, the positive likelihood ratios of 3.5 and greater for the provider documented immunization status for HPV, MCV and Tdap illustrate that patients had a higher probability of needing that immunization if it was documented by a provider. For context, a positive likelihood ratio of 2 and 10 increases the probability of a disease being present by 15% and 45%, respectively.^25,26^ In this study, we defined “disease positive” as being eligible to receive at least one adolescent or influenza immunization based on review of the immunization registry ALERT IIS. The negative likelihood ratios for provider documented immunization status ranged from 0.5-0.9 for all immunizations including when providers documented patients as up-to-date on all immunizations. A LR-of 0.5 and 0.1 decreases the probability of disease or needing the immunization in this study by 15% and 45%, respectively.^25,26^ Provider documentation of immunization status was less predictive of the patient’s true immunization status when the provider documented the patient up-to-date on their immunizations. This highlights the importance of asking about age-specific immunizations needed, rather than asking generically if the patient is up to date, during the initial hospital intake. Alternatively, for patients in states with robust immunization registries, providers may query the registry and confirm the immunizations needed with the patients or families. This strategy may be especially valuable in light of COVID-19 vaccination.

To address the issue of inpatient immunization delivery, multiple strategies have been studied in the pediatric population ranging from identifying those under-immunized and offering immunizations to creating immunization teams.^5,7-9^ Studies evaluating administration of the influenza immunization to hospitalized pediatric patients found that provider reminders, nurse driven screening protocols and immunization ordering tools improved the rates of immunization.^7,27-29^ One study by Moore et al. demonstrated improved immunization rates for the HPV and meningococcal immunizations for hospitalized adolescents by including immunization status in discharge order sets and conducting provider training seminars on the administration of the HPV immunization.^19^ However, accuracy of immunization status documentation by providers remained unchanged despite improvements in the immunizations rates.^19^ Another study by Dempsey et al. evaluated multiple studies to identify strategies to improve adolescent immunization and found promising interventions at the parent and patient level, practice level and population level.^1^ “Practice alerts” embedded into the EMR demonstrated improved HPV immunization rates for the adolescent population in the outpatient setting.^1,30,31^ While these improvements in HPV immunization rates are encouraging, accuracy of provider documentation of immunization status must improve, or states must adopt robust immunization registries, to better immunize this population across outpatient and inpatient settings.

### Limitations

This study has several limitations. This study was conducted at a single urban tertiary care academic children’s hospital with a fairly homogenous population and the results may not be able to be extrapolated to other populations. In addition, 33% of patients in this study either had documentation that the immunization status was unknown, or no documentation of immunization status was present in the admission history and physical, transfer note or discharge summary in the EMR. This represents a large percentage of our study population upon which we were unable to capture provider documentation data of immunization status.

We used the ALERT Immunization Information System (IIS) to determine actual need for immunizations. However, this system may not be 100% accurate and may underrepresent the number of immunizations children receive in the state. ALERT IIS does not currently participate in interstate data exchange so immunizations provided to patients outside the state may not be documented in the system.^13^ In this study, we excluded any patients with zero immunizations in ALERT IIS which may underrepresent the number of unimmunized patients in the population.

## Concluding Summary

This study illustrates that delayed immunizations are prevalent in the hospitalized adolescent population and provider documentation in EMR notes does not accurately reflect their immunization status in the inpatient setting. In our study, 84% of patients were eligible to receive at least one immunization but only one fourth of those eligible were provided an immunization before discharge. This study illustrates the need for better identification and delivery of immunizations to hospitalized adolescents.

## Data Availability

All data produced in the present study are available upon reasonable request to the authors.

## Assistance with study

none.

## Presentation

data presented at trainee research forum.

